# Using unsupervised topic modeling to uncover document hierarchy and latent topics in prostate cancer clinical texts

**DOI:** 10.1101/2024.01.29.24301349

**Authors:** Tuulia Denti, Wei Sun, Shaoxiong Ji, Hans Moen, Oleg Kerro, Antti Rannikko, Pekka Marttinen, Miika Koskinen

## Abstract

**Objective:** Medical records reflect patients’ health status and journey through healthcare services and medical specialties. As a result, document collections contain various text types, even about one patient, which makes medical texts unique. Our aim was to take advantage of the implicit document hierarchy of medical records to evaluate contextualized neural topic modeling as a tool to gain insight into a mixed set of notes related to prostate cancer treatment, i.e., to uncover document types, their contents, and relations.

**Materials and Methods:** We collected clinical text documents from 21,872 prostate cancer patients and organized the documents into a hierarchy using the document metadata. We trained neural topic models without the metadata to index the document collections, performed rigorous numerical evaluations of topic and clustering quality to optimize the topic count, visualized the latent representation of the models, and evaluated the topic clusters with respect to document metadata.

**Results:** Topic clusters reflected the structure of the document hierarchy and provided information about the contents of different text types. The determination of the optimal number of topics required complementary information by topic and clustering quality metrics.

**Discussion:** The topic modeling was found useful in visualizing and indexing large document collections, in providing an understanding of document contents, and in revealing document organization comparable to metadata-based hierarchy.

**Conclusion:** Hospital databases contain masses of text documents, and topic modeling can provide means for analysts and researchers to group documents into discernable and explainable classes.

## 1 Introduction

Clinical notes have long been the primary means of documenting patient care. The amount of text accumulated in electronic health records (EHRs) is often extensive, which calls for effective methods to allow researchers and developers to gain insights into the mass of documents and to make use of the material. Topic modeling [7] refers to a class of unsupervised methods useful for exploring the contents and characteristics of a document collection by assigning topics to texts. Here, we used contextualized topic modeling to uncover structure and topic clusters in clinical text collections.

Clinical notes have unique features compared to texts in many other fields. Notably, texts are written at different stages on the patient pathway and contain distinct text types (classes) with specific contents and vocabularies depending on the medical specialty and document type. In oncology, for example, the number of different text types can be extensive and have hierarchical and temporal relations. Here we show that a latent modeling approach for topic modeling is useful for gaining insights into different document types and their relations in a document collection.

Whereas in recent literature, the validation of topic model performance has been based, e.g., on interpretation of visualizations [32] or topic-word collections [33], we take advantage of document metadata in arranging the documents into a top-down divisive hierarchy, which is also used for topic model evaluation. We demonstrate the close similarity between topic clusters and document hierarchy, as well as the usefulness of topic models in inferring relevant latent topics and themes of the texts. We visualize the hierarchy and perform a rigorous quantitative evaluation of the topic and cluster quality.

## 2 Background

Latent Dirichlet Allocation (LDA) [6] is one of the most popular early topic models to extract latent themes from a collection of free-form text documents. A topic refers to a probability distribution over a fixed vocabulary. The concept is based on the assumption that each document in the collection exhibits a mixture of topics in different proportions. The document-specific topic proportions, topic assignments and topics themselves are the hidden structure inferred from the observed documents [6]. In literature, topic models trained using EHRs have been applied to find relations between topics and variables like genetic cancer mutation test results [9] or the expected duration of mechanical ventilation support [33]. The LDA model has been extended with parameters like clinical note types to improve topic interpretability [33], patients’ diagnoses to discover comorbidity clusters [32], and to account for the change in topic proportions based on, e.g., the document author [27].

As part of electronic health records, clinical notes are connected to several metadata types. Metadata, like report type, age, and diagnosis codes (International Classification of Diseases, ICD) have been used as an additional input to extend the basic LDA model to improve the latent representations [30], to extract meaningful disease topics, and to predict undiagnosed patient phenotypes [18]. Moreover, the metadata has been used as a ground truth to evaluate the quality of clustering [35], and to perform statistical analysis related to patient subgroups extracted with topic modeling [32]. We do not use metadata as an input to the topic modeling. Instead, we use the metadata as a ground truth to validate the topic modeling results, and to show the relation between topics and a hierarchy built with the metadata.

The number of topics depends on the used corpus and is often necessary to be determined prior to the model training. Several measures to select the number of topics have been proposed, such as topic stability [14], model perplexity [36], and cluster quality [16]. The topic modeling results have been evaluated by how well the per-document topic proportions partition the documents into clusters [35] or by measuring the semantic similarity of the topic word collections [21]. The quality of the clusters can be evaluated with internal or external validation measures [19]. The internal validation measures depend only on the information that the clustered data contains and has been evaluated, e.g., with silhouette coefficient [20]. The external validation uses external information or structure, like class labels, for evaluation [34]. Of these external measures, the normalized mutual information and cluster purity have been used to validate the topic modeling results [2; 35; 36; 20]. The quality of the topic word collections can be evaluated with topic coherence, which is defined as semantic similarity of the words within a topic [21] or with topic diversity which is defined with the number of unique words a topic word collection contains [13]. Topic coherence has been measured with, e.g., normalized pointwise mutual information (NPMI) [1; 17; 23; 11]. In this study we present an approach to evaluate topic modeling results based on document cluster quality and the semantic quality of topic word collections. We evaluated the cluster quality with internal and external validation measures, and complementarily the topic word quality with topic coherence and topic diversity metrics. We evaluate the models while changing the number of topics, which gives us an optimal range from which to choose the topic count.

## 3 Materials and Methods

### 3.1 Clinical Notes

In this study we used a collection of clinical notes from 21,872 prostate cancer patients who received treatment in the Hospital District of Helsinki and Uusimaa (HUS), Finland, between December 2004 and February 2021. For these patients, the database contained 1,283,851 clinical notes with a median of 13 (1–299) documents per patient.

The different medical specialties related to the notes and the most common procedures in radiology reports for prostate cancer patients are shown in Figure S1 (supplemental material). We limited surgery and radiation oncology notes to hospital discharges to restrict the size of the dataset. We focused on the selected text categories written during the first two years since the initial diagnosis: full body CT scan reports that measure how widely prostate cancer has spread [3]; prostate MRIs that are used to detect and assess the aggressiveness and local extent of prostate cancer [15]; and bone scans that are used for assessing bone metastasis. All referral notes were omitted, as well as X-ray examinations that are not directly related to prostate cancer diagnostics. The final document collection containing 26,218 clinical texts, which were grouped into four datasets described in Table 1. The process of categorizing, filtering, and labeling the clinical notes is presented in Figure S2 (supplemental material).

**Table 1.**
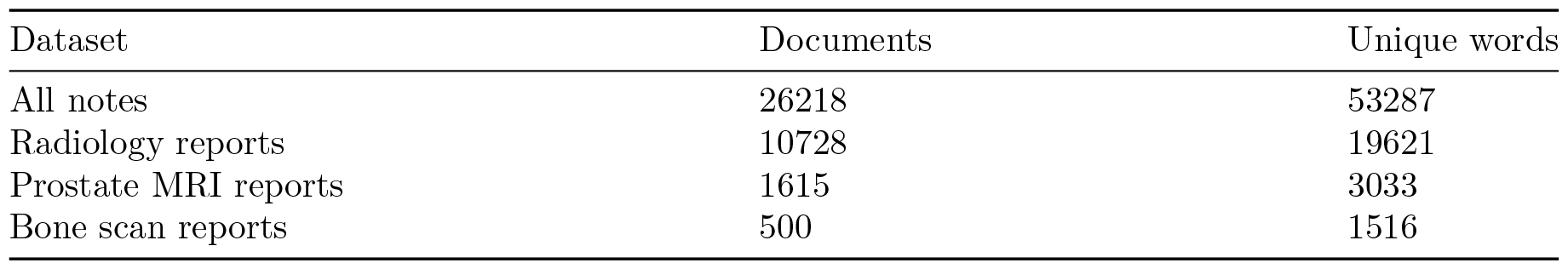
Datasets for topic modeling. The radiology reports are a subset of all notes, and both prostate MRI reports and bone scan reports are subsets of radiology reports. In the Table, word inflections increase the number of unique words.

### 3.2 Contextualized Topic Modeling

We used the neural ZeroShot topic model (ZSTM) by Bianchi et al. [4] to infer topic proportions and vocabulary distributions for each document, and word distributions for each topic. The capability for the model’s zeroshot applications was not utilized. The high-level architecture of the model is described in Figure 1. Contextual sentence representations, generated using Sentence-BERT (SBERT) [25], are functions of the whole text input [12; 22]. SBERT extends the output of a pre-trained BERT with a pooling layer to produce semantically meaningful sentence embeddings [25].

**Figure 1.**
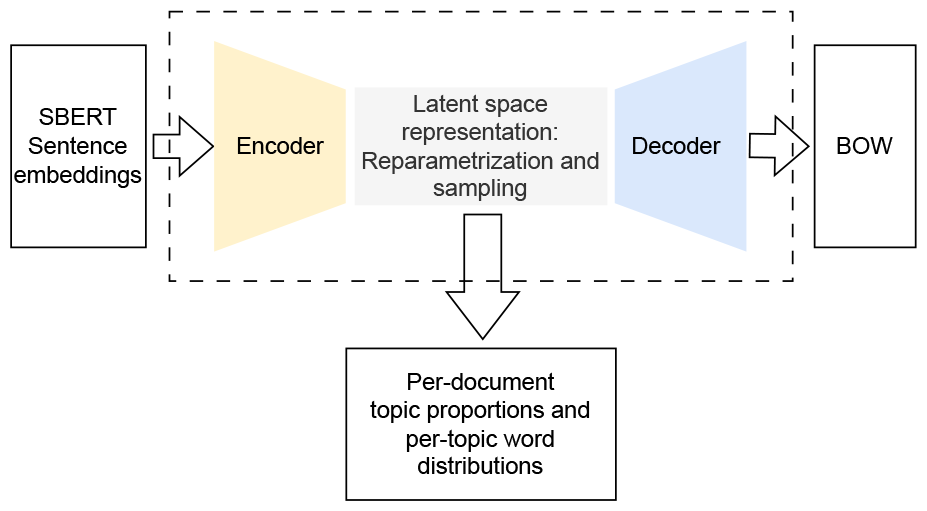
The high-level architecture of the ZSTM [4].

We generated the SBERT embeddings with a Sentence Transformers framework [26] using a pretrained multilingual language model (*paraphrase-multilingual-mpnet-base-v2* ^1^) that allowed analysis of 128 words per text. We trained the ZSTM for 100 epochs and verified the convergence in the training loss separately for all experiments.

The Bag-of-words (BOW) representations of the clinical notes were used to calculate the reconstruction loss during the model training. To create the BOW representations, we removed escape sequences, special characters, numbers, and stop words. For each dataset separately, we calculated using Gensim [24] library the term frequency-inverse document frequency (TF-IDF) values and constructed the dictionary of unique words that was further truncated to 2000 most relevant words [5] using TF-IDF values.

### 3.3 Document Metadata

Each document was accompanied by metadata that was used to assign labels on the clinical notes and to validate the topic modeling result and contained medical specialty, cancer staging system (TNM) codes, and procedures (Nordic Medico-Statistical Committee Classification of Surgical Procedure codes; NCSP) (Figure S1B). The metadata was removed from the documents prior using the documents as training data for the topic models.

The TNM codes that were found meaningful later in our analysis define the primary tumor’s severity and size, and the presence of local and distant metastasis [10]. T-codes from T0 to T4 refer to the tumor’s size and the extent, where T0 denotes no tumor and T4 tumor spreading outside the prostate. M and N-codes denote distant and local metastasis status, respectively. The M-code describes the distant metastasis status. M0 is assigned when there is no distant metastasis, M1 indicates metastasis, and MX represents unclear distant metastasis status. N-code represents whether the tumor has spread to regional lymph nodes, but was omitted since the analyzed reports did not contain N-codes. The labels and the labeling process is described in Figure S2 (supplemental material).

### 3.4 Clustering

We used a topic model-derived clustering method based on the highest probable topic assignment [35]. The most probable topic of each document is set based on the per-document topic proportions, i.e., the topic with the highest proportion in a document is the most probable topic of that document. After each document has been assigned to the most probable topic, the documents are assigned to clusters according to the topics.

### 3.5 Quality Evaluation

The quality assessment of the topic models included the topic quality evaluation based on topic word probabilities and clustering quality evaluation based on per-document topic proportions. For topic quality, we used the Normalized Pointwise Mutual Information (NPMI) [8] score and topic diversity [13]. We defined a high-quality topic as one with a wide collection of cohesively connected words. The NPMI, based on information theory, was used for quantifying the similarity of words in a text and cohesiveness of found topics [1]. This score has been found to associate closely with human-judged coherence value [17]. Given a topic *T* = {*w*_1_, *w*_2_, …, *w*_*N*_ } represented by its top-N most probable words, we calculate the NPMI score [28] of the *i*-th and *j*-th topic word, denoted as

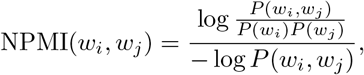

where *P* (*w*_*i*_, *w*_*j*_) is the probability of two words co-occurring in the document and *P* (*w*_*i*_) is the marginal probability of the *i*-th topic word *w*_*i*_. The number of topic words *N* is typically set to 10. For a model that generates *K* topics, the overall NPMI score is an average over topic-specific scores. The range of NPMI is [*−*1, 1], where values close to 1 mean that the topic words appear only together and as such form semantically more cohesive word collections; values close to *−*1 mean that the words occur only separately [8]. The Topic diversity is defined as the fraction of unique words in the 25 most probable words of all topics [13]. Using set notation, this measure can described as

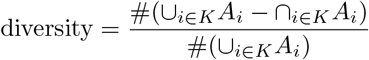

where *K* is the number of topics and *A*_*i*_ is a set of 25 top words in a topic *i*, and # denotes the cardinality of a set. Diversity close to 0 indicates that the topics contain words that are repeated in several topics.

In addition to topic quality, we evaluated the clustering quality using the misclassification rate and silhouette score [29]. We assumed that a high-quality topic clustering relates to distinctly separate groups of documents in the latent space. To calculate the misclassification rate, the documents are assigned into classes based on labels. The possible labels depend on the dataset. For example, the classes for `All notes` and `Radiology reports` datasets are categories and procedures, respectively. The number of classes for each topic is counted, and the highest count becomes the majority class of the topic. The notes that do not share the majority class are labeled misclassified. We calculate the misclassification rate with the fraction of all misclassified notes over the whole set of clustered notes. The silhouette score uses proximities between objects to analyze and evaluate a clustering method. The silhouette score for the whole topic model is calculated using per-document topic proportions as feature vectors and is defined as

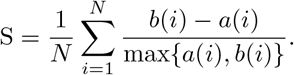

Assume document *i* is assigned to cluster A. The term *a*(*i*) is the average euclidean distance of document *i* to other documents within cluster A. The term *b*(*i*) is the smallest euclidean distance from document *i* to documents not in cluster A. The score range is [-1,1], where values close to one indicate good clustering, and negative values indicate the document is assigned to an incorrect cluster.

### 3.6 Latent Space Visualization

During the quality evaluation, we trained the topic models with a varying number of topics. In order to visualize the latent space of a topic model, we trained a model with the number of topics that yields high NPMI and topic diversity, low misclassification rate, and high silhouette score. The latent space is defined by a document-topic distribution matrix that usually requires dimensionality reduction prior to visualization. We reduced the document-topic distribution matrix dimension with the T-SNE method by [31]. T-SNE converts similarities between data points to joint probabilities and attempts to minimize the Kullback-Leibler divergence between the joint probabilities of the lower-dimensional embeddings and high-dimensional data [31]. The result of the T-SNE method is a set of coordinates in a 2D space each representing a document.

### 3.7 Ethical Aspects

Following national and EU legislation, the study was based on the approval of HUS Helsinki University Hospital (HUS/12199/2022). Data was analyzed in a secure analytics platform (HUS Acamedic) in pseudonymized form.

## 4 Results

### 4.1 Hierarchy Construction

We first obtained document hierarchy as illustrated in Figure 2 using the process and metadata described in Figure S2 (supplemental material)

**Figure 2.**
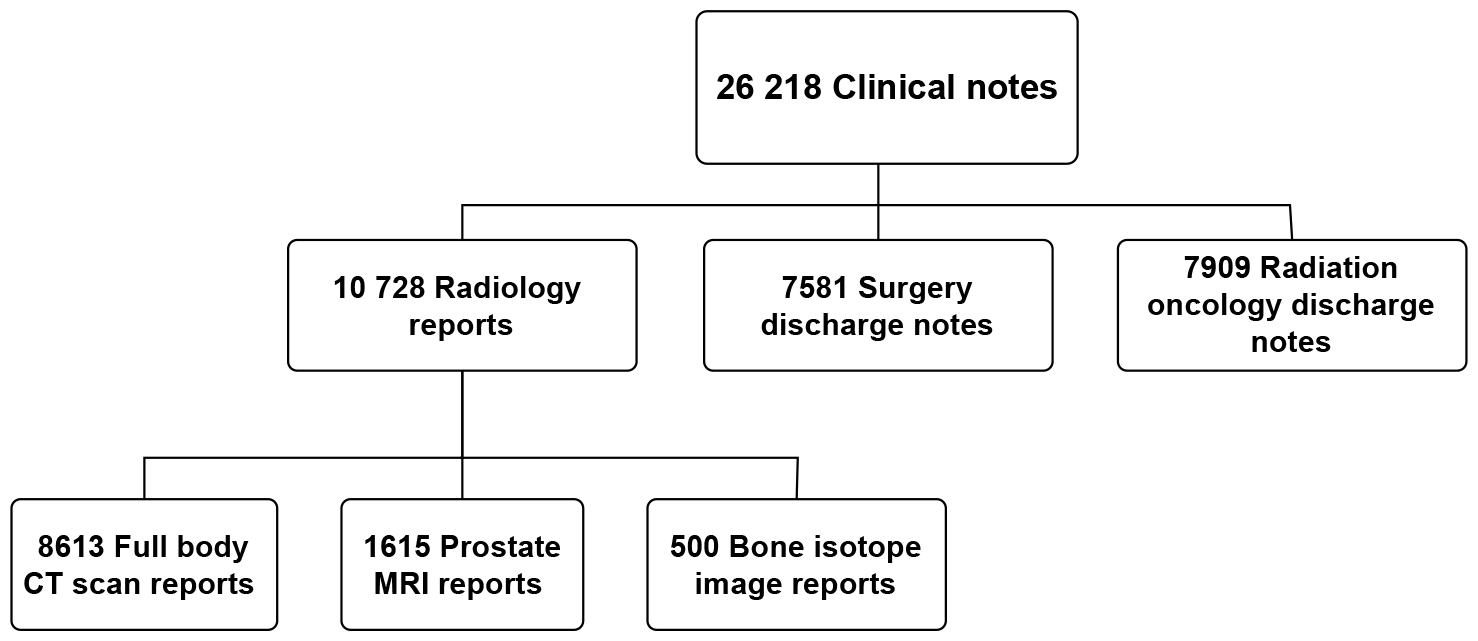
The hierarchy that categorizes the final dataset collection.

### 4.2 Topic and Cluster Quality Evaluation

To find the optimal number of topics based on quality metrics, we trained topic models for each dataset with 2, 4, 6, 8, 10, 12, 16, 20, 40, 60, and 80 topics, three times each. We measured the NPMI, diversity, silhouette score, and misclassification rate and the selected optimal number of topics was a trade-off between the evaluation scores.

Figure 3A shows the results of topic and clustering quality for the All notes dataset. The misclassification rate had lowest values between 10 and 60 topics, while NPMI peaked at 20 with low variation between 10 and 80 topics. The diversity and silhouette scores decreased with the number of topics. Overall the evaluation shows that the optimal topic count is between 6 and 60.

**Figure 3.**
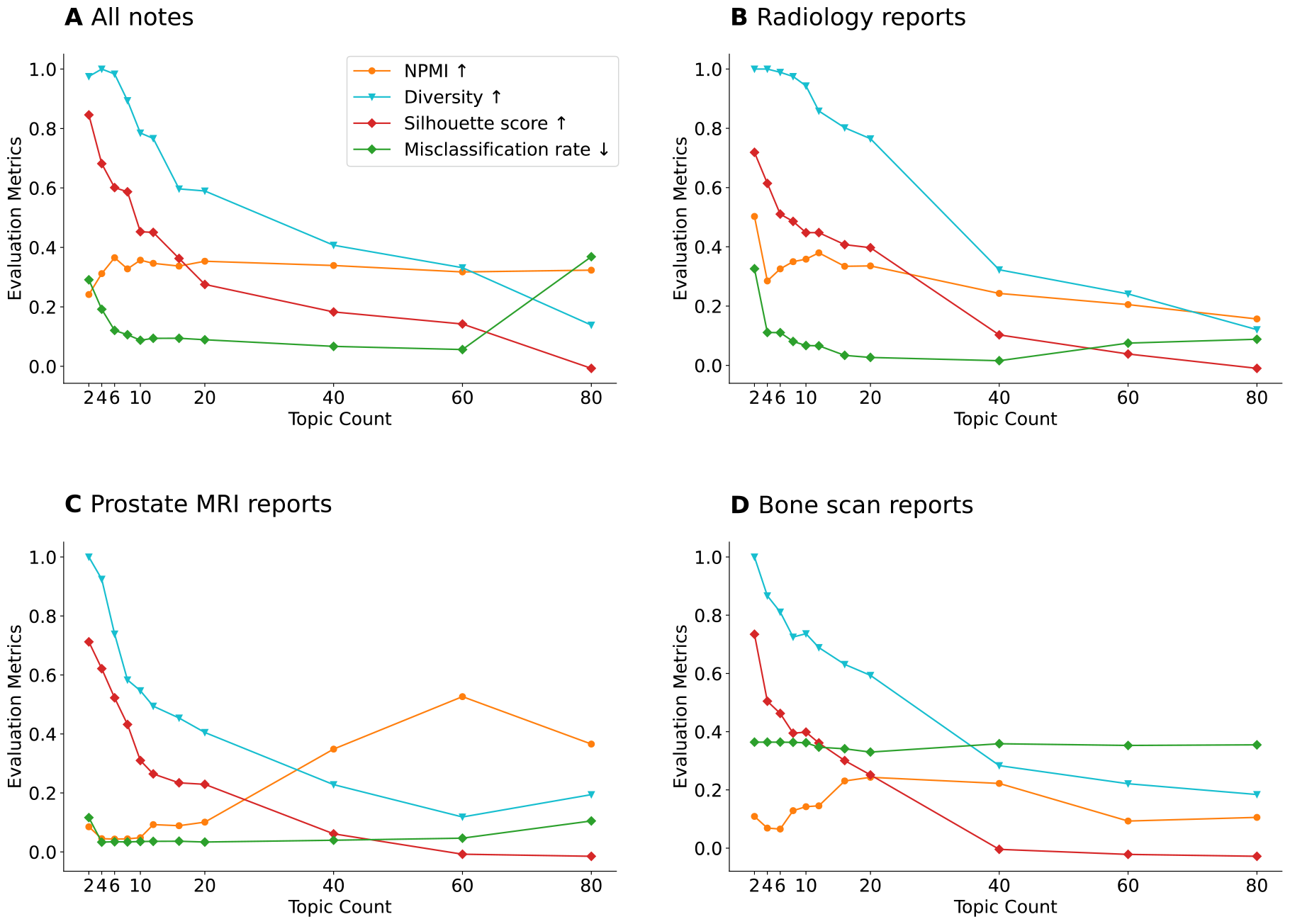
The topic quality evaluated with NPMI and topic diversity, and clustering quality evaluated with silhouette score and misclassification rate. *↑* indicates the higher score, the better performance. *↓* indicates the higher score, the worse performance.

Figure 3B represents the `Radiology reports` dataset and 3C the `Prostate MRI reports` dataset analyzed in similar fashion. Based on these evaluation scores the optimal topic count is between 8 and 20 topics for the `Radiology reports` dataset and between 12 and 20 topics for the `Prostate MRI reports` dataset.

The value of NPMI for the Bone scan report dataset has two almost identical peaks; the first is at 18 topics and the second at 40 topics (Figure 3D). However, the highest diversity or silhouette scores are at two topics. The misclassification rate in the Bone scan report dataset is high and does not vary much. The quality scores are highest for the Bone scan report dataset between 8 and 16 topics.

### 4.3 Latent Space Visualization

The inferred latent space of the topic models was visualized to observe the possible connections between the topics, document clusters, and labels. The number of topics in visualizations were selected based on the quality evaluation, and the number of categories, procedures and T- and M-codes present in each dataset. We aimed to cover all possible latent themes while keeping the number of topics as low as possible and did this by counting the number of labels in dataset and choosing the topic count above this value.

The total number of labels in All notes dataset is 11 (3 medical specialties, 3 procedures, and 5 TNM-codes). Based on the quality evaluation (Figure 3A) the optimal range for the topics is between 6 and 60. Accordingly, the selected number of topics for this dataset was 20. Figure 4A shows that the latent space is split into three main clusters, and these three clusters correspond to the three medical specialties in the ground-truth hierarchy formed with metadata (Figure 2). Furthermore, the area covered by the radiology reports can be divided based on the procedure codes, which is also aligned with the the ground-truth hierarchy.

**Figure 4.**
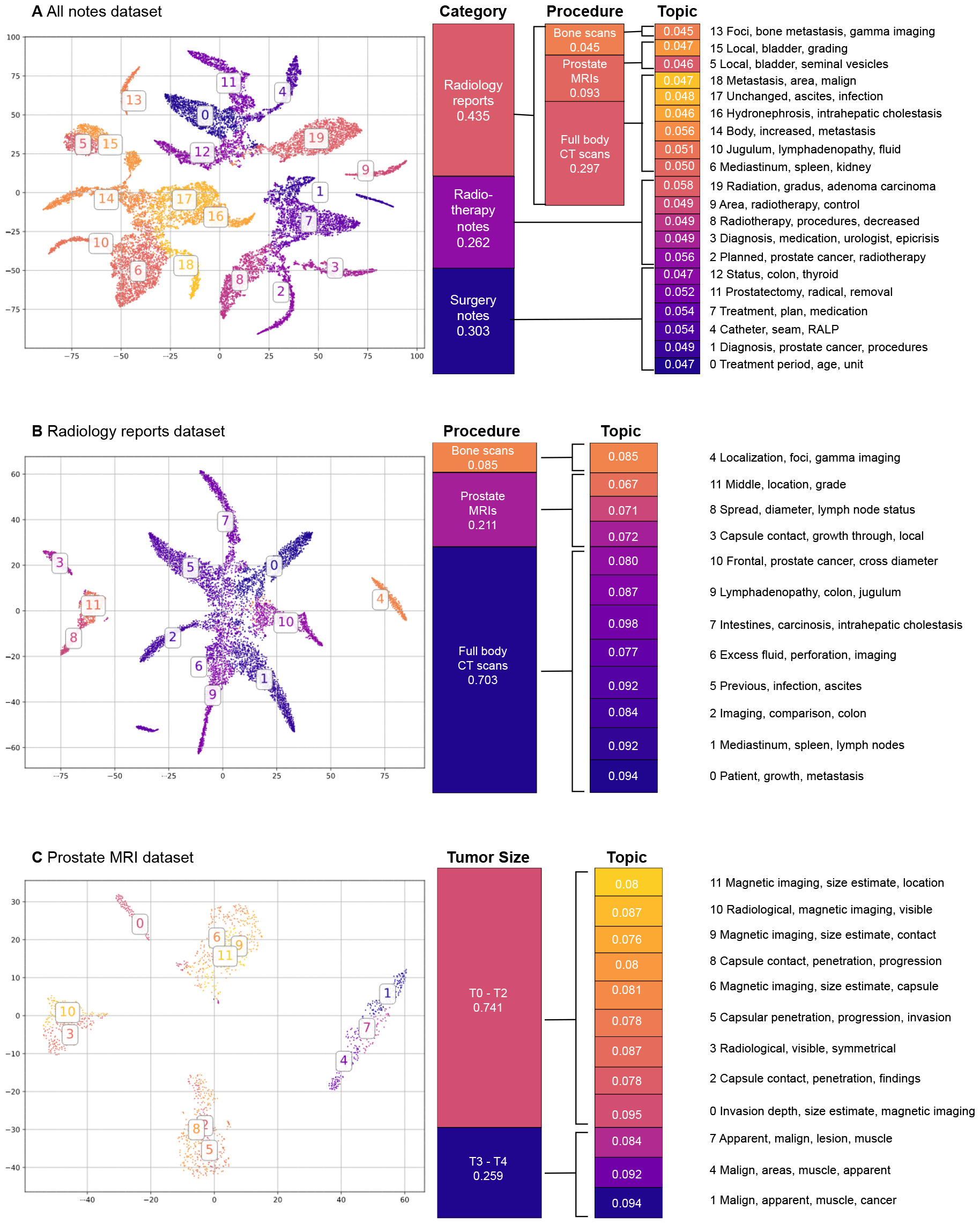
The results of the unsupervised clustering of the documents in datasets containing all texts, radiology reports and prostate MRI reports. The categories and procedures are assigned on topics based on the majority label of the documents within a topic. The topic distributions are the aggregated and normalized topic proportions of the documents. The topics are shown with the most relevant words. The T-SNE projection colors match the topic colors.

The `Radiology reports` dataset contains 8 labels (3 procedures and 5 TNM-codes) and based on the evaluation (Figure 3B) the optimal topic count is between 8 and 20 topics. For the visualization, we chose to use the model trained with 12 topics. Figure 4B of the `Radiology reports` shows that the full-body CT scans cover the largest area, while prostate MRIs and bone scans occupy smaller, clearly separate areas. This corresponds to the lowest level of the ground-truth hierarchy in Figure 2.

For the `Prostate MRI reports dataset`, the optimal topic range is between 12 and 20 topics (Figure 3C), and the dataset contains 3 labels (a single procedure with two T-codes). Since the dataset and the vocabulary size are relatively small, we accepted a lower NPMI score and visualized the latent space of the `Prostate MRI reports` dataset with 12 topics. The visualization for the `Prostate MRI reports` dataset in Figure 4C shows clearly separated clusters. The T0-T2 codes cover 4 clusters that are separate from the cluster containing documents with T3-or T4-codes. This shows that the the unsupervised analysis using the topic models was able to divide the documents into clusters based on tumor size.

The optimal topic range for the `Bone scan report` dataset is between 8 and 16 topics (Figure 3D) and the dataset contains 4 labels (a single procedure with 3 M-codes). Based on this, we trained the model with 8 topics. Figure S3 (supplementary material) for the `Bone scan report` dataset shows that the M0-code dominates the topics. We did not find any connections between the topics and bone metastasis in the bone scan reports likely due to small sample size, the imbalanced label distribution, and the cluster assignment method used.

## 5 Discussion

We found that contextual topic modeling can serve as an unsupervised approach for providing meaningful insights into a large collection of clinical notes with respect to content, types of notes, and relations between the notes. Even though the metadata was not part of the training data, the latent representation by the ZSTM model reflected the ground-truth document hierarchy that was constructed using the metadata on medical specialties and procedures.

The clusters in the latent space representation of the three largest datasets showed a connection between the hierarchy classes represented by the labels and the topics. While some clusters overlapped, the topics were grouped in a way that matches the top levels of the divisive hierarchy. The visualization of the `All notes` dataset showed that the documents and topics can be arranged according to the hierarchy: first based on category, then based on the procedure. Additionally, the latent representations and topic proportions of the `Prostate MRI reports` dataset showed a connection between the learned topics and the size and severity of the cancer tumor, for which the ground-truth was encoded in the T-codes. A connection with M-codes was not evident.

The highest topic probability based clustering is a simple and computationally inexpensive method that connected the clusters and topics directly. The method ignored part of the information about the per-document topic distributions. When a document contained topics in equal proportions, the most probable topic did not represent the document entirely, and the clustering was not representing the documents. Other clustering methods, like k-means, could be used to partition the documents, but the clear connection between the labels and topics would be lost.

We considered several evaluation metrics to find the optimal topic count for each dataset. Our topic and cluster quality evaluation results showed that some of the metrics contradict each other and that there was no single optimal number of topics and we determined an optimal range for the topic count. The NPMI is biased against low-frequency word collocations [8]. For the datasets with small vocabularies many of the word collocations became high-frequency when the number of topics increased which lead to an increase of NPMI value. The topic diversity favored the lowest topic numbers, since the probability of overlapping word collections decreased. However, some overlap in topic word collections was expected and acceptable. The silhouette score favored the lowest topic numbers and decreased when the number of topics increased. This is due to the clusters beginning to overlap when the number of them increased. The misclassification rate depended on the most probable topic each document was assigned to and it increased with both low and high topic counts. This indicated that the complexity of the model was not high enough to explain the data with the lower number of topics and lead to overfitting with highest number of topics.

## 6 Conclusion

Our study showcased the potential of applying cutting-edge contextualized text representation and neural topic models to process unlabeled clinical notes in the healthcare information system by applying the contextualized topic model to clinical text analytics in real-world data collected from prostate cancer patients. We presented an approach to utilize metadata derived labels to create a top-down divisive hierarchy to categorize a large set of documents and showed a connection between this ground-truth 5hierarchy and the learned document classes through visualization. Additionally, we found that topic quality evaluation metrics like NPMI and topic diversity are insufficient to determine the optimal number of topics in a topic model on their own, but the optimal number can be found as their combination. Our empirical results can provide a practical means for analysts and researchers to gain insight on a large mixed set of clinical documents. External signals, such as metadata, can be used to guide the first steps of the analysis as a computationally light and efficient method to explore the content of the clinical texts.

## Acknowledgments

This work was supported by the Academy of Finland (Flagship programme: Finnish Center for Artificial Intelligence FCAI, and grants 315896, 336033) and EU H2020 (grant 101016775). This work started as a Master thesis (without peer review) in the Master’s Programme in Computer, Communication and Information Sciences at Aalto University, jointly with HUS Helsinki University Hospital.

## Author contributions statement

T.D. programmed the software and visualized the results. T.D. wrote the original draft. All authors reviewed and edited the manuscript, and analyzed and interpreted the data. T.D., W.S., S.J., H.M., P.M., and M.K. analyzed the results. S.J. and M.K. supervised the project. O.K. curated the data.

P.M. acquired funding. P.M. and M.K. administered and planned the project.

## Data availability

Due to national legislation, restrictions apply to the availability of clinical data at the individual level, which were used with the permission of HUS Helsinki University Hospital.

## Informed Consent Statement

No ethical permission was required according to the Finnish Medical Research Act for the secondary use of medical records. Following national and EU legislation, the study was based on the approval of HUS Helsinki University Hospital (permission HUS/12199/2022)

## Supplementary Materials

**Figure S1.**
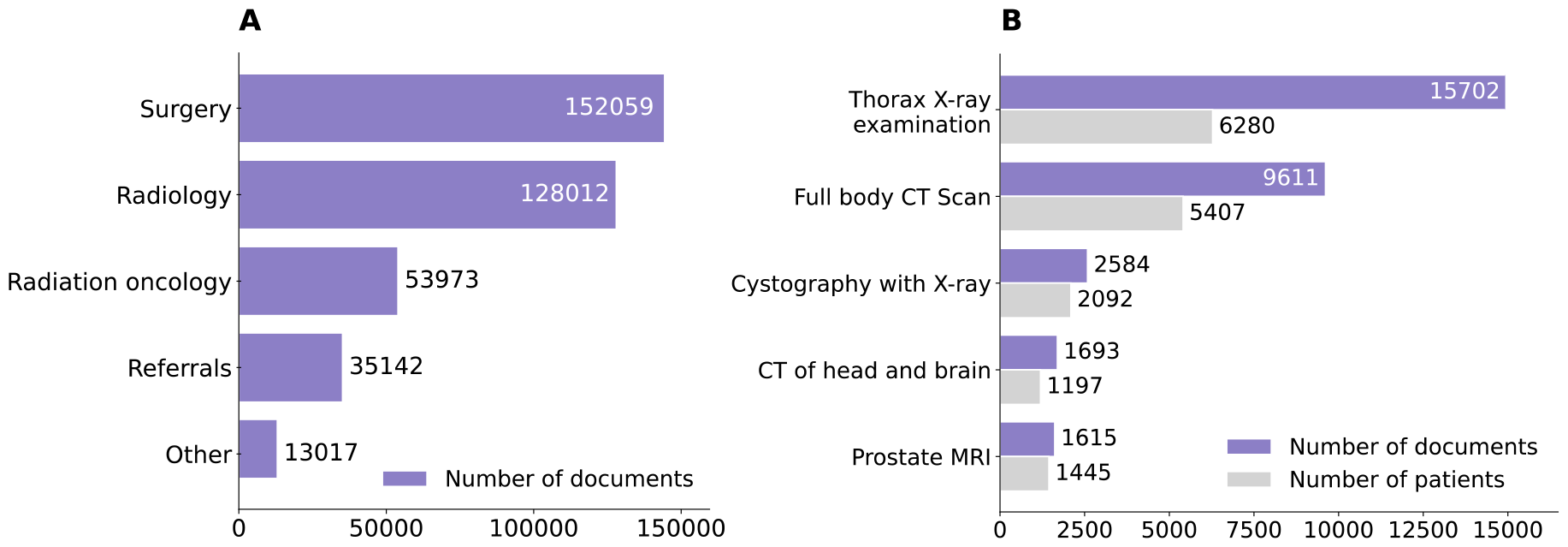
A. Most common medical specialties in the clinical note collection. The category ‘other’ contains notes from, e.g., clinical physiology, dental surgery, and internal medicine. B. Most common procedures (Nordic Medico-Statistical Committee Classification of Surgical Procedure codes; NCSP) in the clinical note collection.

**Figure S2.**
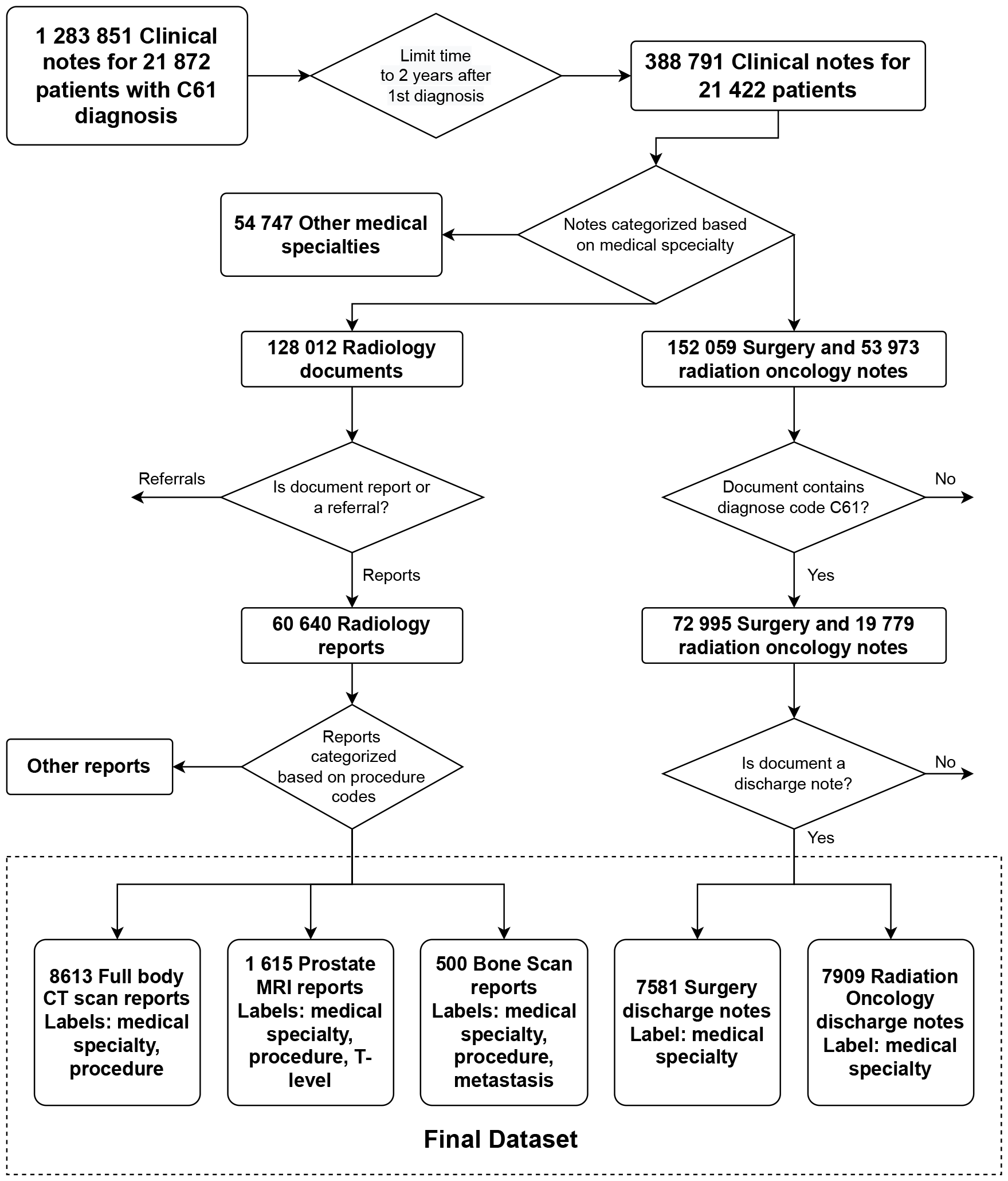
The categorization and selection process of the documents to form the final dataset

**Figure S3.**
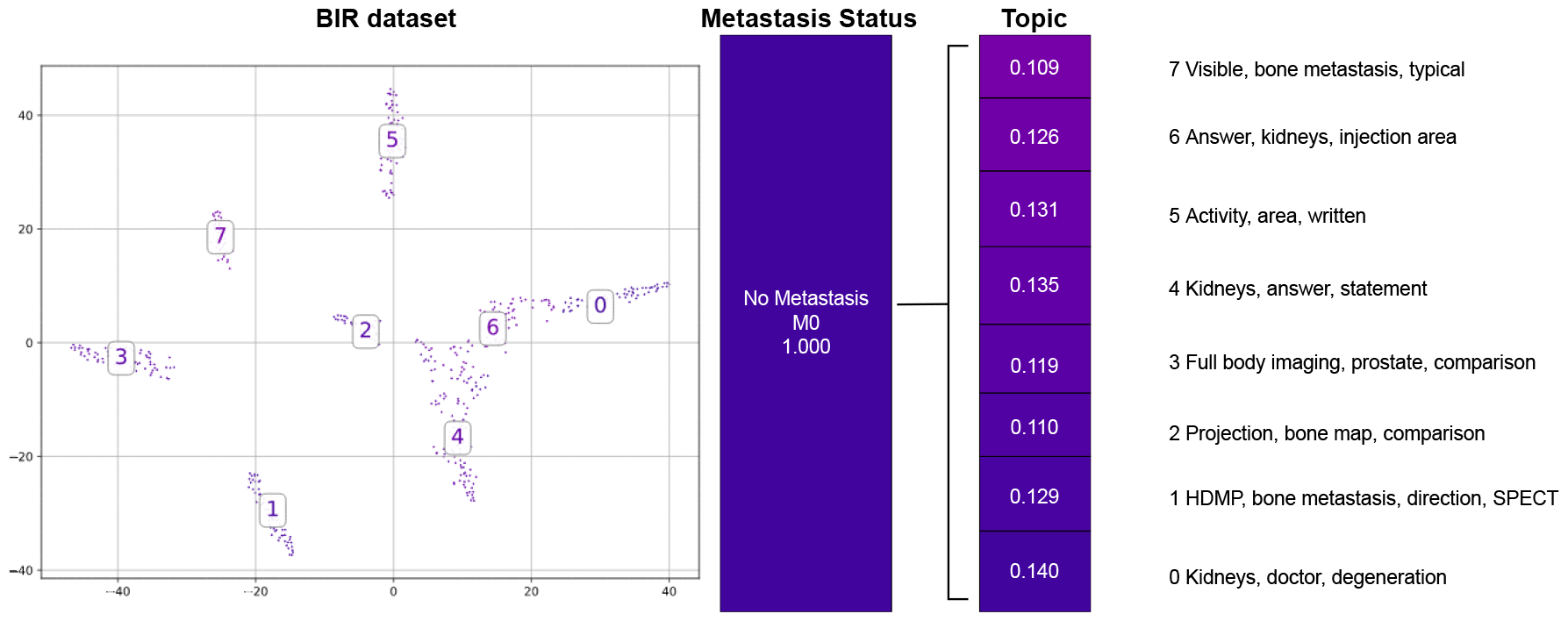
The clustering of the documents in the `Bone scan report` dataset. The aggregated and normalized topic proportions for M-codes and Topics.

## Footnotes

1 https://huggingface.co/sentence-transformers/paraphrase-multilingual-mpnet-base-v2

